# Outcomes, Trends, and Healthcare Disparities in Patients Hospitalized with Chronic Limb Threatening Ischemia

**DOI:** 10.1101/2023.05.25.23290559

**Authors:** Christian Torres, Francisco Ujueta, Everett Rogers, Amre Ghazzal, Radleigh Santos, Christian Koelbl, Esteban Escolar, Gervasio A. Lamas, Sahil A Parikh, Nirat Beohar

## Abstract

**Background:** Chronic limb threatening ischemia (CLTI) is the most severe form of peripheral artery disease (PAD) and is associated with poor patient outcomes and increased healthcare costs. While racial, socioeconomic, and other healthcare disparities are widely recognized to influence the management of CLTI, the extent of the role they play is still an area of intense investigation.

**Methods:** We analyzed data from the National Inpatient Sample (NIS) to identify all patients ≥18 years of age admitted with a primary diagnosis of CLTI from 2016 to 2019. Descriptive statistics were used to summarize patient baseline characteristics (age, gender, race, comorbidities, socioeconomic status, and procedural rates). Logistic regression models and temporal trends were used to determine predictors of major amputation and MACE, as well as in CLTI admissions during the 4-year study period, major amputation, endovascular intervention, and peripheral bypass further divided into racial cohorts.

**Results:** A total of 121,087,650 patients were hospitalized from 2016 to 2019 of which 4,707,657 (3.9%) hospitalized for CLTI. The mean age of patients admitted with CLTI was 60 ± 17 years. A majority were male (57.8%, p<0.001), and White (72.0%, p<0.001). They were more likely to be socioeconomically disadvantaged (32.8% with median household income 0-25^th^ percentile, p<0.001). Risk for hospitalization for CLTI varied inversely with increasing household income. During the hospitalization, 32.4% had invasive angiography, 0.6% had peripheral computed tomography angiogram (CTA), 3.3% underwent angioplasty, 1.6% peripheral bypass, and major amputation occurred in 9.2%. Black patients had the highest risk for amputation, followed by Native American and Hispanic patients. White patients made up the greatest percentage of CLTI admissions, but were not at increased risk for amputation. Asian and Pacific Islander patients were the only racial group at decreased risk for amputation. Temporal trends during the 4-year period revealed the strongest predictors of MACE were diabetes and a history of peripheral angioplasty or peripheral bypass. Overall, there was a 6.7% increase in hospitalizations, a 14.1% increase in peripheral angioplasty rates, and an 8.4% decrease in peripheral bypass rates for CLTI during the 4-year study period. There was a reduction in above the knee amputation rates for all racial cohorts except for Native Americans (23.5% increase) during the study period. There was a 26.4% total increase in below the knee amputation rates.

**Conclusion:** Despite increased awareness of health disparities, poor outcomes resulting from CLTI (such as amputation) continue to disproportionately affect racial and socioeconomic minority groups. Revascularization and amputations during hospital admission for CLTI is increasing, driven by peripheral angioplasty and BKA, respectively.

## Background

Peripheral arterial disease (PAD) is estimated to affect 236.6 million people over 25 years of age worldwide (1). The most severe stage of PAD, chronic limb threatening ischemia (CLTI), leads to increased cardiovascular mortality, lower extremity amputations, and, at the societal level, increased healthcare expenditures. Diabetes mellitus (DM), cigarette use, hypertension (HTN), hyperlipidemia, obesity, and sedentary lifestyle have been recognized as risk factors for CLTI (2). Ethnic and racial disparities, and socioeconomic status also play a role in the outcomes of CLTI, with previous retrospective studies demonstrating greater odds of lower extremity amputation among Black compared with White patients(3–5). Previous studies have demonstrated that individuals with PAD as a whole are medically undertreated. For example, approximately 60% are treated with HMG-CoA reductase inhibitor (statin) therapy(3,6). However, research over the last several decades has suggested that the group predominantly affected by the underutilization of medical and revascularization-based therapies are Black patients(7). It has been repeatedly shown that Black patients are less likely to undergo limb salvage therapy (i.e., lower extremity revascularization), an important treatment in CLTI, likely resulting in the increased rates of lower extremity amputation seen in this group(8). Health disparities are defined by preventable markers of health differences in a population group, such as ethnicity, race, sex, education level, geographic location, and household income (9). Although much focus has been placed in traditional risk factors such as hypertension, diabetes, obesity, and tobacco cigarettes, other risk factors should be explored to continue to decrease the incidence and outcomes of CLTI. The 2021 American Heart Association (AHA) Statement of Peripheral Artery Disease identified non-conventional risk factors for PAD, which included urban areas and environmental contaminants (ex. air pollution, metals contaminants)(10).

The purpose of the present study is to use the Nationwide Inpatient Sample (NIS) database to explore factors that contribute to racial and ethnic disparities in the treatment of CLTI. In addressing possible factors of disparities in CLTI we hope to answer: (1) What are the baseline characteristics of patients admitted to the hospital for CLTI; (2) Are any of these characteristics able to help predict the risk of amputation and/or major adverse cardiovascular events (MACE), (3) Specifically, how do race and household income affect patient’s risk for amputation or MACE; (4) How are amputation and revascularization rates for CLTI changing over time for the CLTI population as a whole, and for racial subgroups?

## Methods

### Data Source

Data were obtained from the NIS database from 2016 through 2019. The NIS is a publicly available database that is part of the Healthcare Cost and Utilization Project (HCUP) and sponsored by the Agency for Healthcare Research and Quality (AHRQ) created with federal, state, and industry partnerships (1). It is the largest publicly available all-payer inpatient healthcare database in the United States (US), yielding national estimates of inpatient hospital outcomes. It consists of a stratified sample of discharges from all hospitals in HCUP and it equals approximately 20% of all discharges in US hospitals. Each hospitalization event included in the database represents a unique observation conformed by one primary discharge diagnosis and 29 secondary diagnoses during the index hospitalization. Using weights (DISCWT) provided by the NIS database, it estimates more than 35 million hospitalizations per year nationally(11). Since the database contains de-identified patient information, the study was deemed exempt from the need for Institutional Review Board (IRB) approval by our institution.

### Study Population

Using International Classification of Diseases, Tenth Edition, Clinical Modification (ICD-10-CM) codes we identified all patients older than 18 years of age that were admitted between 2016 and 2019 with a primary diagnosis of CLTI (ICD-10^th^ CM I7022, I7023, I7024, I7025, I7026, I7032, I7033, I7034, I7035, I7036, I7042, I7043, I7044, I7045, I7046, I7052, I7053, 7054, I7055, I7056, I7062, I7063, I7064, I7065, I7066, I7072, I7073, I7074, I7075, I7076).

### Patient and hospital variables

Baseline characteristics, socioeconomic information, hospital characteristics, and demographics were obtained utilizing variables provided by the NIS database(11). Socioeconomic data included median household income per quartiles. Demographics consisted of age, gender, and race. Relevant clinical variables were obtained using separate ICD-10 codes and Elixhauser clinical comorbidities as defined by the AHRQ (2). Additionally, diagnostic tests and procedures like computed tomography angiography (CTA), invasive peripheral angiography, peripheral angioplasty, peripheral bypass, above and below the knee amputation rates were obtained using the ICD-10-PCS codes. A detailed table of the ICD-10-CM and ICD-10-PCS codes used to identify patient comorbidities and procedures are provided in Appendix Table 1.

**Table 1.**
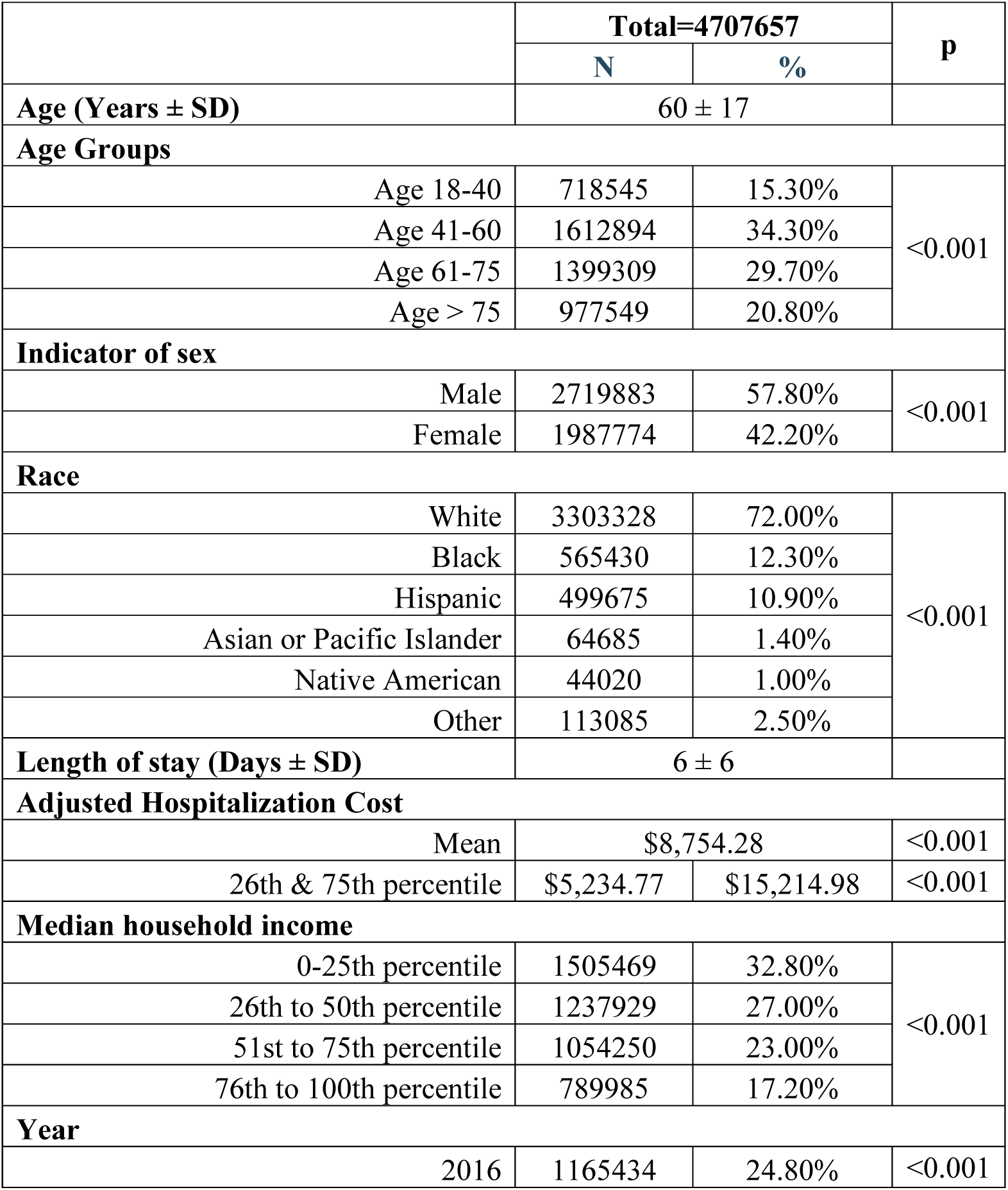
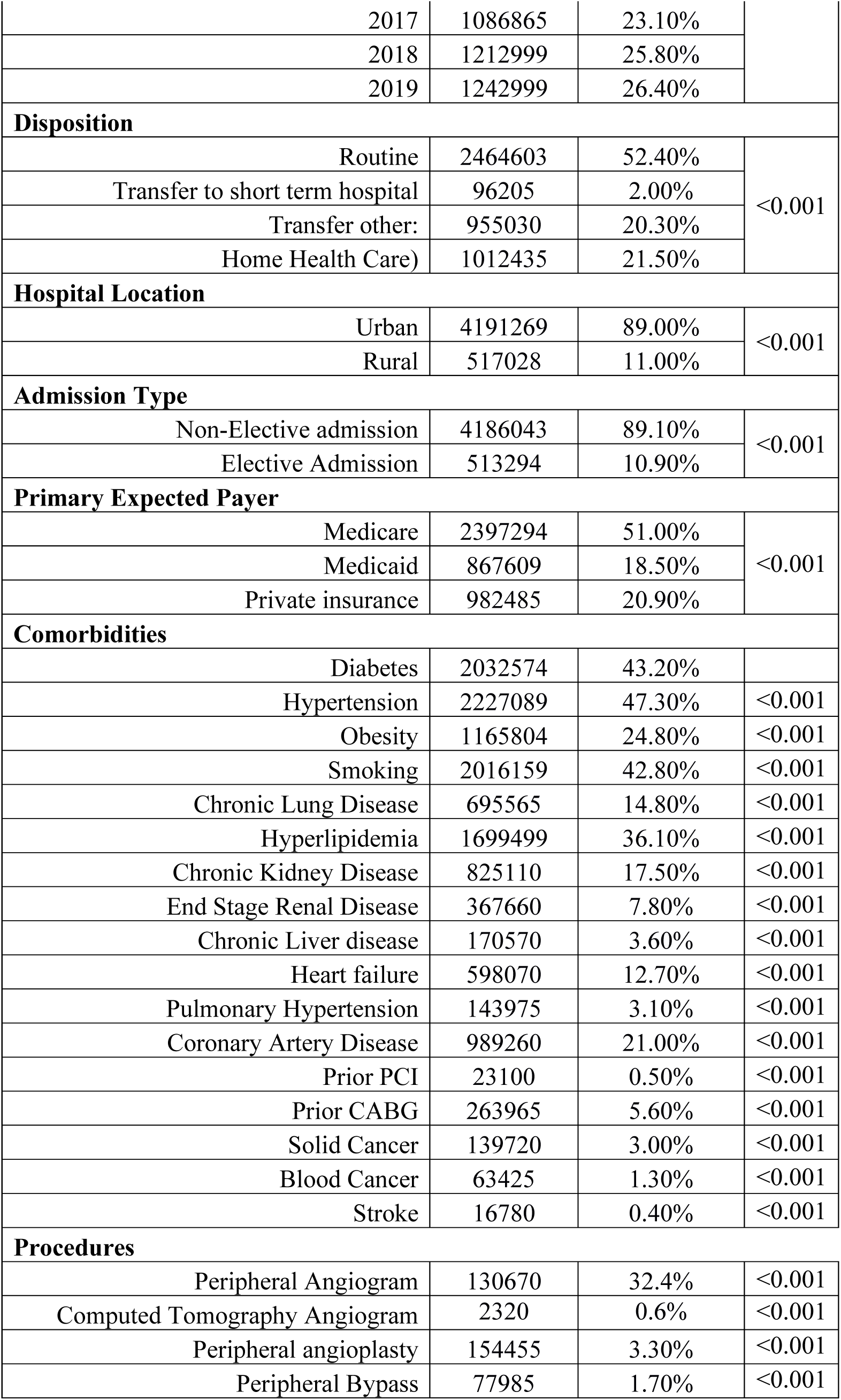
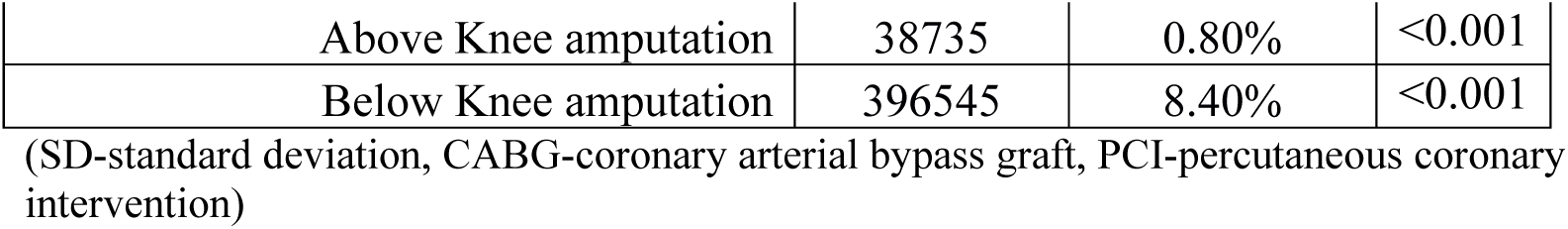
Baseline Characteristics of Patients Admitted to the Hospital with Chronic Limb Threatening Ischemia (2016-2019).

|  | Total=4707657 |  | p |
| --- | --- | --- | --- |
|  | N | % |  |
| Age (Years ± SD) | 60 ± 17 |  |  |
| Age Groups |  |  |  |
| Age 18-40 | 718545 | 15.30% | <0.001 |
| Age 41-60 | 1612894 | 34.30% |  |
| Age 61-75 | 1399309 | 29.70% |  |
| Age > 75 | 977549 | 20.80% |  |
| Indicator of sex |  |  |  |
| Male | 2719883 | 57.80% | <0.001 |
| Female | 1987774 | 42.20% |  |
| Race |  |  |  |
| White | 3303328 | 72.00% | <0.001 |
| Black | 565430 | 12.30% |  |
| Hispanic | 499675 | 10.90% |  |
| Asian or Pacific Islander | 64685 | 1.40% |  |
| Native American | 44020 | 1.00% |  |
| Other | 113085 | 2.50% |  |
| Length of stay (Days ± SD) | 6 ± 6 |  |  |
| Adjusted Hospitalization Cost |  |  |  |
| Mean | \$8,754.28 | | <0.001 |
| 26th & 75th percentile | \$5,234.77 | \$15,214.98 | <0.001 |
| Median household income |  |  |  |
| 0-25th percentile | 1505469 | 32.80% | <0.001 |
| 26th to 50th percentile | 1237929 | 27.00% |  |
| 51st to 75th percentile | 1054250 | 23.00% |  |
| 76th to 100th percentile | 789985 | 17.20% |  |
| Year |  |  |  |
| 2016 | 1165434 | 24.80% | <0.001 |
| 2017 | 1086865 | 23.10% |  |
| 2018 | 1212999 | 25.80% |  |
| 2019 | 1242999 | 26.40% |  |
| Disposition |  |  |  |
| Routine | 2464603 | 52.40% | <0.001 |
| Transfer to short term hospital | 96205 | 2.00% |  |
| Transfer other: | 955030 | 20.30% |  |
| Home Health Care) | 1012435 | 21.50% |  |
| Hospital Location |  |  |  |
| Urban | 4191269 | 89.00% | <0.001 |
| Rural | 517028 | 11.00% |  |
| Admission Type |  |  |  |
| Non-Elective admission | 4186043 | 89.10% | <0.001 |
| Elective Admission | 513294 | 10.90% |  |
| Primary Expected Payer |  |  |  |
| Medicare | 2397294 | 51.00% | <0.001 |
| Medicaid | 867609 | 18.50% |  |
| Private insurance | 982485 | 20.90% |  |
| Comorbidities |  |  |  |
| Diabetes | 2032574 | 43.20% |  |
| Hypertension | 2227089 | 47.30% | <0.001 |
| Obesity | 1165804 | 24.80% | <0.001 |
| Smoking | 2016159 | 42.80% | <0.001 |
| Chronic Lung Disease | 695565 | 14.80% | <0.001 |
| Hyperlipidemia | 1699499 | 36.10% | <0.001 |
| Chronic Kidney Disease | 825110 | 17.50% | <0.001 |
| End Stage Renal Disease | 367660 | 7.80% | <0.001 |
| Chronic Liver disease | 170570 | 3.60% | <0.001 |
| Heart failure | 598070 | 12.70% | <0.001 |
| Pulmonary Hypertension | 143975 | 3.10% | <0.001 |
| Coronary Artery Disease | 989260 | 21.00% | <0.001 |
| Prior PCI | 23100 | 0.50% | <0.001 |
| Prior CABG | 263965 | 5.60% | <0.001 |
| Solid Cancer | 139720 | 3.00% | <0.001 |
| Blood Cancer | 63425 | 1.30% | <0.001 |
| Stroke | 16780 | 0.40% | <0.001 |
| Procedures |  |  |  |
| Peripheral Angiogram | 130670 | 32.4% | <0.001 |
| Computed Tomography Angiogram | 2320 | 0.6% | <0.001 |
| Peripheral angioplasty | 154455 | 3.30% | <0.001 |
| Peripheral Bypass | 77985 | 1.70% | <0.001 |
| Above Knee amputation | 38735 | 0.80% | <0.001 |
| Below Knee amputation | 396545 | 8.40% | <0.001 |

(SD-standard deviation, CABG-coronary arterial bypass graft, PCI-percutaneous coronary intervention)

### Outcomes Definition

We primarily sought to determine predictors of adverse outcomes (i.e., major amputation (major adverse limb events (MALE) usually defined as: either major amputation of the revascularized limb and/or reintervention on the revascularized segment) and major adverse cardiac events (MACE)) among patients hospitalized for CLTI, with a focus on evaluating for race/ethnic disparities, and socioeconomic status during the study period. As secondary outcomes, we evaluated patient baseline and hospital characteristics (including hospital location (Urban/Rural), hospitalization costs, and length of stay (LOS)). In addition, temporal trends in hospitalizations, amputations, and revascularization rates for patients with CLTI were assessed.

### Statistical analysis

Discharge weight provided by the NIS was applied to unweighted data to infer national estimates in accordance with HCUP regulations (12). Weighted data were used in all analyses. We used descriptive statistics to summarize the distribution of baseline parameters and hospital characteristics. Continuous variables were expressed as means or medians (costs), standard deviations, and 25th-75th percentiles respectively, while categorical variables were expressed as absolute values with percentages. To ascertain differences between variables, we used the Pearson chi-square, Fisher’s exact test, independent t-test, and Mann-Whitney U test as appropriate. Binary-logistic regression was used to explore the association between CLTI and inpatient outcomes. First, we performed a univariable analysis; variables that were significantly associated with our outcomes (p <0.2) were included in our multivariable model to adjust for potential confounders. To estimate the temporal changes during the study period, we used time-series regression models. All statistical analyses were conducted using SPSS Statistics 27.0 (IBM Corp., Armonk, New York). All p-values were two-sided and statistical significance was defined as p<0.05.

## Results

A total of 121,087,650 patients were admitted for any cause during the 48-month study period between January 2016 and December 2019, of which 4,707,657 (3.9%) were admitted with CLTI listed in the top three diagnoses. Admissions subdivided by race were 78,939,896 (65.2%) White, 17,814,860 (14.7%) Black, 13,044,454 (10.8%) Hispanic, 3,235,038 (2.7%) Asian, and 753,290 (.6%) Native American.

### Baseline Characteristics

The mean age of patients admitted with CLTI was 60 ± 17 years of age (standard deviation). Patients with CLTI were more likely to be male (57.8% vs. 42.2%, p<0.001). Both White (72.0% vs. 65.2%, p<0.001) and Native Americans (1.0% vs 0.6%, p<0.001) made up a greater proportion of CLTI admissions compared to admissions for any cause. Conversely, Black (12.3% vs 14.7%, p<0.001) and Asian or Pacific Islander patients (1.4% vs 2.7%, p<0.001) made up a smaller proportion of CLTI admissions compared to admissions for any cause. Although still statistically significant, rates of admission for Hispanic patients was roughly similar in both the CLTI and any cause groups (10.9% vs 10.8%, p<0.001). (Table 1).

When divided by median household income quartiles, patients with CLTI were more likely to be economically disadvantaged. The highest percentage of patients (32.8%) were in the lowest income quartile (i.e., median household income in the 0-25th percentile). The risk for hospitalization with CLTI decreased as household income increased. The most common comorbidities among patients with CLTI included hypertension (47.3%), diabetes (43.2%), smoking (42.8%), hyperlipidemia (36.1%) and obesity (24.8%).

In terms of procedures, 32.4% of patients admitted with CLTI underwent invasive angiography, 0.6% had peripheral CTA, 3.3% peripheral angioplasty, and 1.6% underwent peripheral bypass during index hospitalization. Major amputation occurred in 9.2% of patients; 0.8% of patients underwent above the knee amputation (AKA) and 8.4% underwent below the knee amputation (BKA) during index hospitalization.

### Predictors of Amputation

The age group with the largest number of patients (between 41 and 60-years-old) had the highest risk for amputation (adjusted odds ratio (aOR) 1.41, 95% CI: 1.40-1.43, p<0.001) (Table 2). Females were significantly less likely to undergo amputation (aOR 0.59, 95% CI: 0.58-0.59, p<0.001). Among racial cohorts, Black patients were at the highest risk for undergoing amputation during hospitalization (aOR 1.37, 95% CI: 1.34-1.39, p<0.001), followed by Native American (aOR 1.37, 95% CI: 1.32-1.42, p<0.001) and Hispanic (aOR 1.12, 95% CI: 1.10-1.14, p<0.001) patients. White patients were not at increased risk for amputation (aOR 1.00, 95% CI: 0.99-1.02, p=0.860), and Asian and Pacific Islander patients were the only racial group at decreased risk for amputation (aOR 0.80, 95% CI: 0.77-0.83, p<0.001) (Table 3).

**Table 2.** Multivariate Predictors of Amputation in Patients Admitted with Chronic Limb Threatening Ischemia (2016-2019)

| Variables | aOR | 95% CI | p-value |
| --- | --- | --- | --- |
| Age 18 - 40 | 0.75 | 0.74-0.76 | p<0.001 |
| Age 41 - 60 | 1.41 | 1.40-1.43 | p<0.001 |
| Age 61 - 75 | 1.30 | 1.29-1.32 | p<0.001 |
| Female | 0.59 | 0.58-0.59 | p<0.001 |
| African American | 1.37 | 1.34-1.39 | p<0.001 |
| White | 0.86 | 0.99-1.02 | p=0.86 |
| Hispanic | 1.12 | 1.10-1.14 | p<0.001 |
| Asian | 0.80 | 0.77-0.83 | p<0.001 |
| Native American | 1.37 | 1.32-1.42 | p<0.001 |
| Urban | 0.77 | 0.76-0.78 | p<0.001 |
| Diabetes | 4.15 | 4.12-4.19 | p<0.001 |
| Hypertension | 1.13 | 1.12-1.13 | p<0.001 |
| Obesity | 0.66 | 0.65-0.67 | p<0.001 |
| Smoking | 1.08 | 1.07-1.08 | p<0.001 |
| Chronic Lung Disease | 0.79 | 0.78-0.79 | p<0.001 |
| Hyperlipidemia | 1.15 | 1.15-1.16 | p<0.001 |
| Chronic Kidney Disease | 1.23 | 1.22-1.24 | p<0.001 |
| End Stage Renal Disease | 1.35 | 1.34-1.37 | p<0.001 |
| Chronic Liver Disease | 0.57 | 0.56-0.59 | p<0.001 |
| Heart Failure | 0.76 | 0.75-0.77 | p<0.001 |
| Atrial Arrhythmias | 0.89 | 0.88-0.90 | p<0.001 |
| Coronary Artery Disease | 1.17 | 1.16-1.18 | p<0.001 |
| Prior PCI | 0.94 | 0.90-0.98 | p<0.001 |
| Stroke | 0.62 | 0.58-0.66 | p<0.001 |
| Prior CABG | 1.04 | 1.03-1.05 | p<0.001 |
| 0 – 25<sup>th</sup> Percentile | 1.25 | 1.23-1.28 | p<0.001 |
| 26 – 50<sup>th</sup> Percentile | 1.23 | 1.20-1.26 | p<0.001 |
| 51-75<sup>th</sup> Percentile | 1.18 | 1.15-1.21 | p<0.001 |
| 76 – 100<sup>th</sup> Percentile | 1.05 | 1.02-1.07 | p<0.001 |
| Peripheral Angioplasty | 3.10 | 3.06-3.14 | p<0.001 |
| Peripheral Bypass | 1.61 | 1.58-1.64 | p<0.001 |

(aOR- adjusted odds ratio, CI- confidence interval, CABG- coronary artery bypass grafting, PCI- percutaneous coronary intervention)

**Table 3.** Multivariate Predictors of Major Adverse Cardiovascular Events in Patients Admitted with Chronic Limb Threatening Ischemia (2016-2019).

| Variables | aOR | 95% CI | p-value |
| --- | --- | --- | --- |
| Age 18 - 40 | 0.12 | 0.11-0.13 | <0.001 |
| Age 41 - 60 | 0.41 | 0.40-0.42 | <0.001 |
| Age 61 - 75 | 0.7 | 0.69-0.71 | <0.001 |
| Female | 1.02 | 1.00-1.04 | p=0.012 |
| African American | 0.89 | 0.86-0.93 | <0.001 |
| White | 0.93 | 0.90-0.96 | <0.001 |
| Hispanic | 0.8 | 0.76-0.83 | <0.001 |
| Asian | 1.35 | 1.26-1.44 | <0.001 |
| 0 - 25th Percentile | 1.07 | 1.01-1.13 | p=0.013 |
| 26 - 50th Percentile | 1.03 | 0.98-1.09 | p=0.237 |
| 51-75th Percentile | 0.95 | 0.90-1.00 | p=0.049 |
| 76 - 100th Percentile | 0.9 | 0.85-0.95 | <0.001 |
| Native American | 0.99 | 0.90-1.08 | p=0.769 |
| Urban Hospital | 0.98 | 0.95-1.00 | p=0.127 |
| Diabetes | 1.07 | 1.06-1.09 | <0.001 |
| Hypertension | 0.89 | 0.87-0.89 | <0.001 |
| Obesity | 0.83 | 0.81-0.85 | <0.001 |
| Smoking | 0.87 | 0.86-0.89 | <0.001 |
| Chronic Lung Disease | 0.98 | 0.96-0.99 | p=0.017 |
| Hyperlipidemia | 1.04 | 1.03-1.06 | <0.001 |
| Chronic Kidney Disease | 1.03 | 1.02-1.05 | <0.001 |
| End Stage Renal Disease | 1.28 | 1.25-1.31 | <0.001 |
| Chronic Liver Disease | 1.26 | 1.21-1.31 | <0.001 |
| Heart Failure | 1.61 | 1.57-1.64 | <0.001 |
| Atrial Arrhythmias | 1.26 | 1.23-1.28 | <0.001 |
| Coronary Artery Disease | 2.46 | 2.41-2.50 | <0.001 |
| Prior PCI | 0.92 | 0.86-0.99 | p=0.038 |
| Prior CABG | 0.86 | 0.84-0.88 | <0.001 |
| Peripheral Angioplasty | 1.14 | 1.10-1.18 | <0.001 |
| Peripheral Bypass | 1.25 | 1.19-1.31 | <0.001 |
| Above Knee Amputation | 1.7 | 1.60-1.79 | <0.001 |
| Below Knee Amputation | 0.59 | 0.57-0.60 | <0.001 |

(aOR- adjusted odds ratio, CI- confidence interval, CABG- coronary artery bypass grafting, PCI- percutaneous coronary intervention)

Just as patients from lower socioeconomic households were more likely to be hospitalized with CLTI, they were also more likely to undergo amputation during index hospitalization (aOR 1.25, 95% CI: 1.23-1.28, p<0.001). We found this risk to be sequentially stratified. In other words, progressively lower median household income quartile was associated with increased risk for amputation (Table 3).

While the most common baseline characteristics of patients admitted with CLTI were well recognized atherogenic risk factors, the top three predictors of amputation included DM (aOR 4.15, 95% CI: 4.12-4.19, p<0.001), history of peripheral angioplasty (aOR 3.10, 95% CI: 3.06-3.14, p<0.001), and history of peripheral bypass (aOR 1.61, 95% CI: 1.58-1.64, p<0.001) (Table 3).

### Predictors of MACE

In contrast to predictors of amputation, age was not a significant predictor of in-hospital MACE risk (Table 3, Figure 2). Asian and Pacific Islanders were the only racial cohort at increased risk for MACE during hospitalization (aOR 1.35, 95% CI: 1.27-1.44, p<0001). The risk for MACE was again sequentially stratified among the different socioeconomic groups, with the lowest socioeconomic households being the only stratum at increased risk (aOR 1.07, 95% CI: 1.02-1.13, p=0.013).

**Figure 1.**
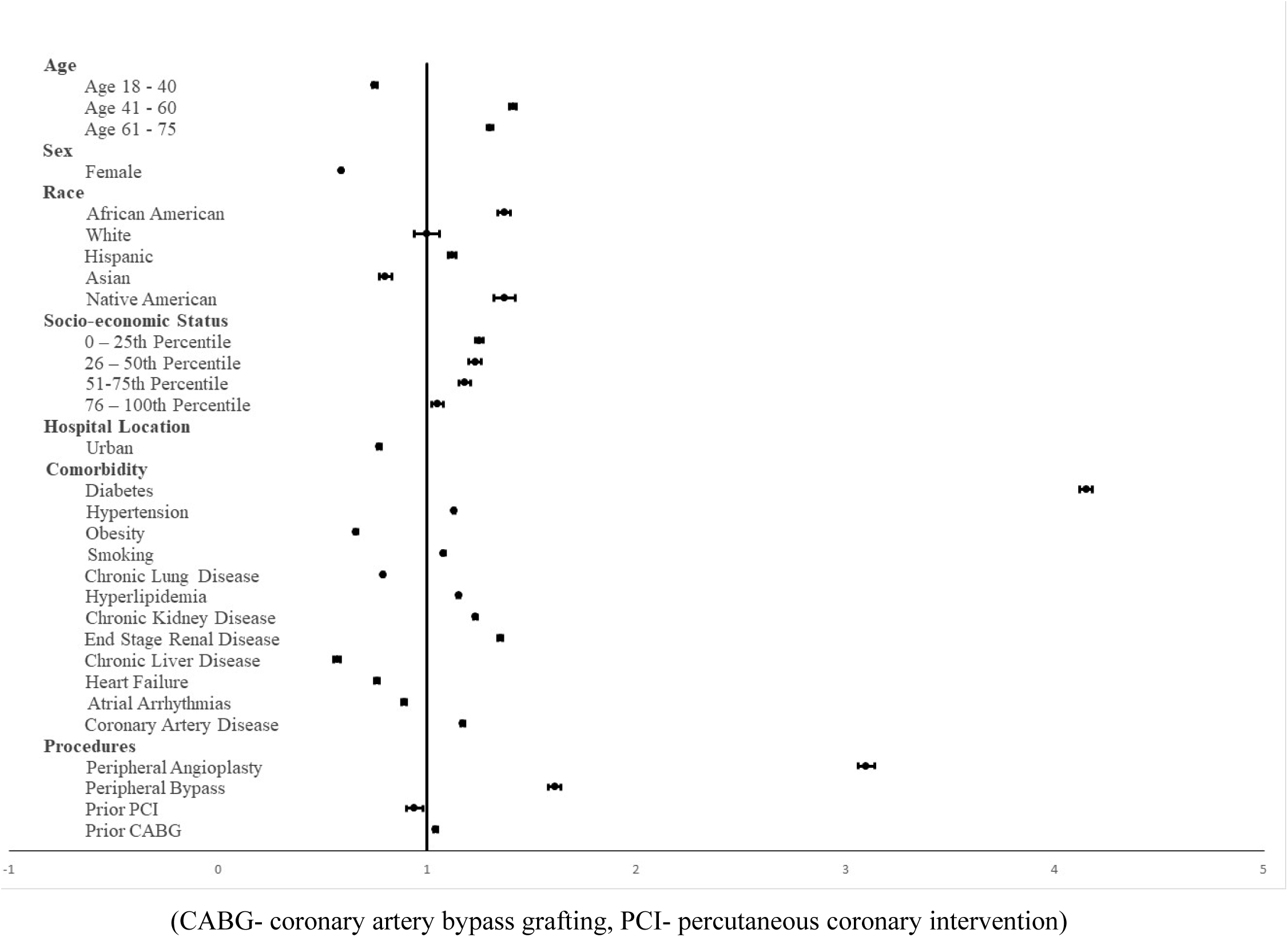
Forest Plot of Predictors of Amputation in Patients Admitted with Chronic Limb Threatening Ischemia (2016-2019)

**Figure 2.**
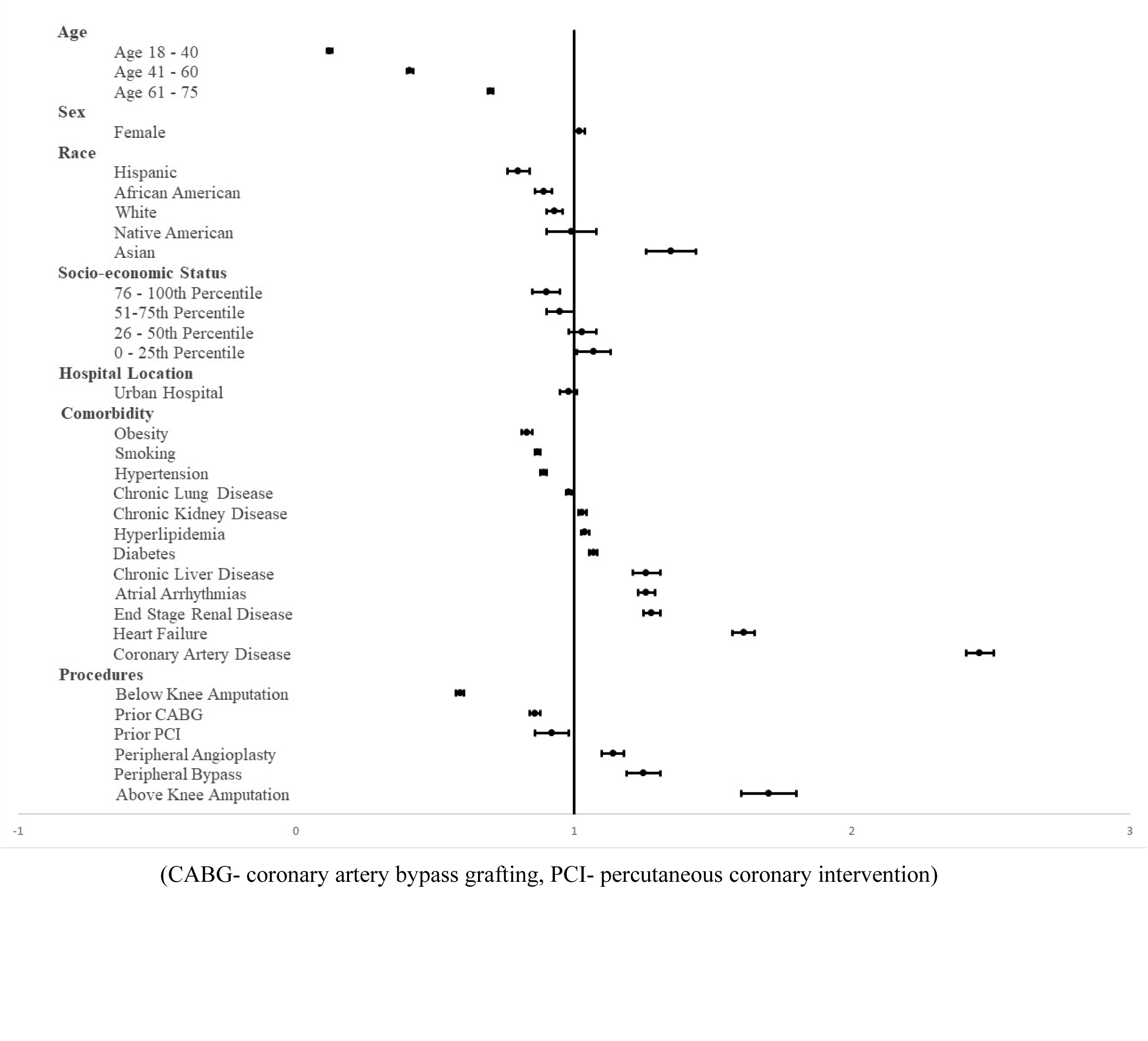
Forest Plot of Predictors of Major Adverse Cardiovascular Events in Patients Admitted with Chronic Limb Threatening Ischemia (2016-2019).

Overall, the top three predictors of MACE in patients admitted with CLTI were coronary artery disease (aOR 2.46, 95% CI: 2.41-2.50, p<0.001), history of AKA (aOR 1.70, 95% CI: 1.61-1.79, p<0.001), and heart failure (aOR 1.61, 95% CI: 1.57-1.64, p<0.001) (Table 2).

### Temporal Trends

In spite of an initial downward trend in admissions for CLTI from 2016 to 2017, there was an overall 6.7% increase in CLTI admissions during the study period (Figure 3). Major amputations increased by 22.7% during the study period, driven by a 26.4% increase in BKA. The increase in BKA was seen in all racial cohorts. White patients saw the largest increase in BKA in terms of raw numbers (16,340 more procedures performed in 2019 compared to 2016), while Native Americans saw the largest percent increase (51.96% more procedures performed in 2019 compared to 2016). AKA decreased by 8.71% during the study period, driven largely by a reduction in AKA among Black patients. A reduction in AKA was seen in all racial cohorts, except for Native Americans who experienced a 23.5% increase in AKA, although the total number of AKA performed in this population was very low.

**Figure 3.**
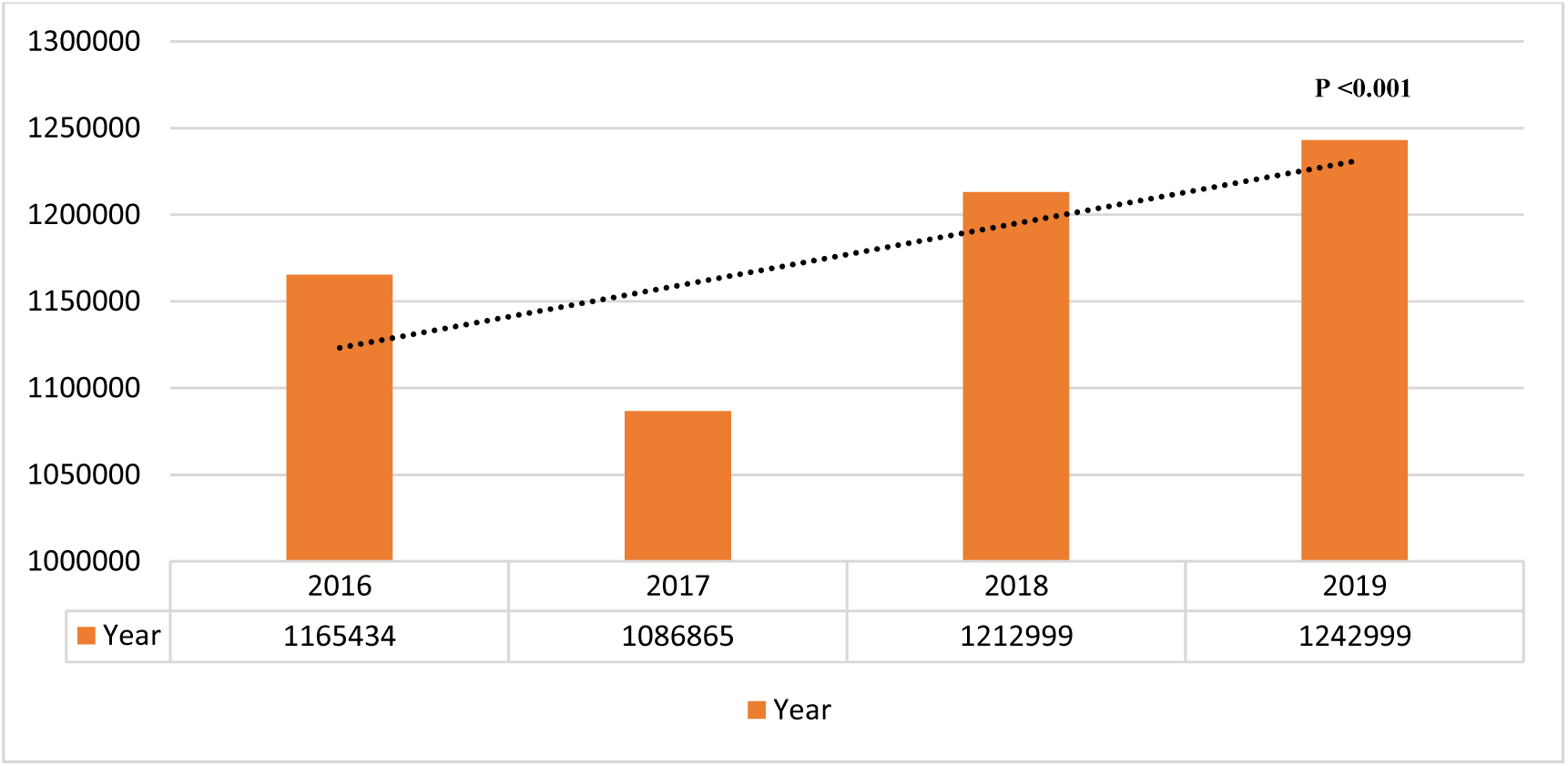
Yearly Trends of Patients Admitted with Chronic Limb Threatening Ischemia (2016-2019).

## Discussion

In a large population-based study the following were the principal findings: 1) A majority of patients were White males and were more likely to be socioeconomically disadvantaged. 2) The risk for hospitalization for CLTI varied inversely with increasing household income. 3) During hospitalization, catheter based or surgical revascularization was infrequent, and major amputation was three times more likely than revascularization. 4) Black patients had the highest risk for amputation, followed by Native American and Hispanic patients. White patients were not at increased risk for amputation, and Asian and Pacific Islanders were the only racial group at decreased risk for amputation. 5) The strongest predictors of MACE were DM and a history of prior peripheral revascularization. 6) There was an overall increase in hospitalizations and peripheral angioplasty rates, but a decrease in peripheral bypass rates for CLTI during the study period. 7) There was a reduction in above the knee amputation rates for all racial cohorts except for Native Americans, but an increase in below the knee amputation rates for all racial subgroups.

The present study demonstrated that household income and ethnic disparities are risk factors in lower extremity amputation and in-hospital mortality in patients admitted for PAD-CLTI. In particular, Black and Native American racial cohorts appeared to be impacted the greatest. Known risk factors such as DM and a history of lower extremity revascularization have the highest predictive value for lower extremity amputation; patients with DM having 4x greater risk, odds ratio (aOR 4.15 (4.12-4.19). It was also suggested that patients undergoing peripheral angioplasty have a 3x higher risk of lower extremity amputation, (aOR 3.10 (3.06-3.14). Admissions for CLTI are on the rise, which may be due to increased awareness in identifying this patient population or under management of the disease with subsequent progression to CLTI and eventually amputation. The majority of CLTI admissions occur in urban compared to suburban hospitals, which could be due to the increased population density or increased individual exposure to risk factors such as air pollution (such as PM_2.5_) and metal contaminants (ex. cadmium and lead, found in water, food, and air pollution)(13–15).

Black patients are known to have a 2-to 3-fold higher prevalence of PAD(16,17) compared with White individuals(18,19). These racial differences may be due to the higher prevalence of traditional risk factors in Black communities, such as DM, obesity, and smoking. Furthermore, previous studies have shown that Black communities have lower access to proven cigarette smoking cessation treatments, making them more vulnerable to relapse and increasing their lifetime exposure to cigarette smoke(20,21). Despite the increased prevalence of atherogenic risk factors in the Black population, epidemiological studies have not implicated these factors as the sole reason for the increased prevalence of PAD and disproportionately poor outcomes seen in this group(22). This observation is supported by an analysis of the National Health and Nutritional Examination Survey from 1999 to 2000, which included over 2000 participants over 40 years of age. It demonstrated that Black individuals have an approximately 3x higher associated prevalence of PAD compared to White individuals (OR 2.83), even after accounting for smoking status, DM, body mass index, glomerular filtration rate, and hypercholesteremia(23). Thus, although traditional therapies and interventions have focused on decreasing conventional risk factors of PAD, non-traditional risk factors such as environmental pollutants, race, socio-economic status, and certain comorbidities (including depression, elevated lipoprotein (a), and CKD) may be playing a role in the increased prevalence and likely outcomes in PAD-CLTI.

Our results are consistent with prior studies revealing low household income being a risk factor for worse outcomes and lower extremity amputations(3–5). This study suggests that Americans in the lowest income quartile account for almost one-third of admissions for CLTI, and have the highest risk for amputation and MACE. In addition, the lowest quartile income are also at a 25% increased risk of major non-traumatic amputations. Although PAD-CLTI as a whole continues to be underrecognized and undertreated by the medical community, Black patients are disproportionately undertreated with optimal medical therapy compared to other racial cohorts. A study using electronic health records from the University of Michigan Health system demonstrated that Black patients were significantly less likely to be prescribed a statin for primary prevention, despite having a higher atherosclerotic risk score(24). A similar study from Emory University revealed a 66.7% statin prescription rate for Black patients with PAD, versus 73.4%(p<0.001) for all other racial groups(25). Antiplatelets have also been shown to be underutilized in this population. In one small single center study from Gober et al., all patients who were inappropriately discharged without antiplatelets were Black, and half were female(26). In addition, several studies have demonstrated that Black patients are less likely to be offered limb salvage therapies such as revascularization (27–30). Thus, this raises the question, is it provider bias, patient access to optimal medical therapy, or a system failure in providing optimal medical management to Black patients with PAD-CLTI that results in the increased prevalence of CLTI, and eventual lower extremity amputations seen in this population?

Social determinants of health do not only include household income or access to healthcare as suggested previously, but also housing. It has been suggested that biological mechanisms such as a proinflammatory state and endothelial oxidative stress likely play a role in the disparity(31), but one must look further. In 2021, the AHA identified air pollution (ex., PM_2.5_) and heavy metals (such as lead and cadmium) as non-traditional risk factors for PAD(15). The majority of Black individuals are located in the urban cities of the United States, closer to major roadways and industrialization, which exposes them to daily environmental insults, which we now know increases cardiovascular mortality. Environmental contaminants such as cadmium and lead are two well-known cardiovascular vasculotoxins, which cause endothelial dysfunction, increase reactive oxygen species and inflammatory factors, resulting in atherosclerosis. Lead production increased during the turn of the 20^th^ century, with extensive industrial and public use, including lead gasoline and household paint, in addition to being found in tobacco cigarettes. Interestingly, although smoking cessation decreases the risk of PAD-CLTI, a study has shown that it takes 30 years for an individual’s risk to reach that of a never smoker(32), and the half-life of cadmium and lead in the human body is approximately 30 years. Cadmium has the strongest association with PAD. Our group previously published a study in patients with coronary artery disease, demonstrating urinary cadmium was lowest in those without PAD, higher in individuals with PAD, and highest in CLTI individuals(14). Thus, although our focus has helped decrease conventional risk factors in PAD-CLTI, our continued exposure to non-traditional risk factors may be a modifiable means to curb the increasing incidence. There is a great need for public health interventions through legislation to control these risk factors and to minimize the inciting exposure of air pollution and metal contamination in the water, soil, and food(33).

### Native Americans

Our study also revealed an increased rate of major amputations in Native Americans in patients admitted with CLTI. The Strong Heart Study, a population-based cohort study in 12 Native American communities recruited between 1989-1991, has demonstrated in various published studies the interaction between environmental contaminants and increased cardiovascular disease (13). The AHA in a recent statement identified metal contaminants, particularly lead and cadmium, as cardiovascular risk factors in this ethnic population (13,34). The source of this metal contamination is likely water wells and dietary sources, which heightens the importance of safe accessible drinking water(35,36).

### Study Limitations

Similar to any retrospectively collected data set, the study may include biases related to possible erroneous diagnosis or diagnostic misclassification or under-reporting. These errors may have been more pronounced in the study considering the NIS relies on ICD-10-CM codes. Proper diagnosis of CLTI by general clinicians can be difficult, though the noted increase in CLTI hospitalization may in part be related to increased recognition. Some factors are not able to be objectively captured by the NIS database. For example, some patient characteristics, such as race, are self-reported, leading to potential reporting bias. Furthermore, the NIS only captures procedures that were performed during the index admission. Therefore, if a patient admitted with CLTI were discharged and underwent a procedure in the outpatient setting, our data would not capture the event and lead to underreporting. This is of importance as peripheral vascular interventions are being done in outpatient base catheterization labs. Similarly, if a patient were discharged from the hospital and suffered a MALE or MACE sometime later, our data is not able to associate that event with the index CLTI admission.

## Conclusions

The rates of CLTI increased during our 4-year review of the NIS database. While AKA and peripheral bypass rates are decreasing, the number of BKA and peripheral angioplasties performed for CLTI are significantly increasing. It is important for future studies to continue to monitor these trends, particularly in the context of recently published data from the Best Endovascular Versus Best Surgical Therapy in Patients with CLTI (BEST-CLI) trial suggesting that surgical revascularization may lead to more favorable outcomes in with CLTI for whom both surgical and endovascular intervention is feasible. DM and a history of previous revascularization procedures are the most important predictors of increasing in-hospital lower extremity amputations. Among racial and socioeconomic cohorts, individuals in the lowest quartile of household income and Black and Native American patients have the highest risk of non-traumatic major lower extremity amputations. We must look at modifiable non-traditional risk factors such as environmental contaminants, reimbursements, wound care, obesity education, socio-economic impediments to medical care, and certain comorbidities (including depression, elevated lipoprotein (a), and CKD) as an additional source to continue decreasing our rates of cardiovascular events in CLTI. In addition, we need to continue to expand and improve the delivery of accessible clinical care to all individuals, regardless of race, sex, or socioeconomic status. Additional clinical trials and community studies are needed to further understand the racial disparities and potential interventions needed to decrease the risk of negative limb outcomes and improve health equity.

## Data Availability

The authors confirm that the data supporting the findings of this study are available within the article and its supplementary materials.

## Supplemental Material

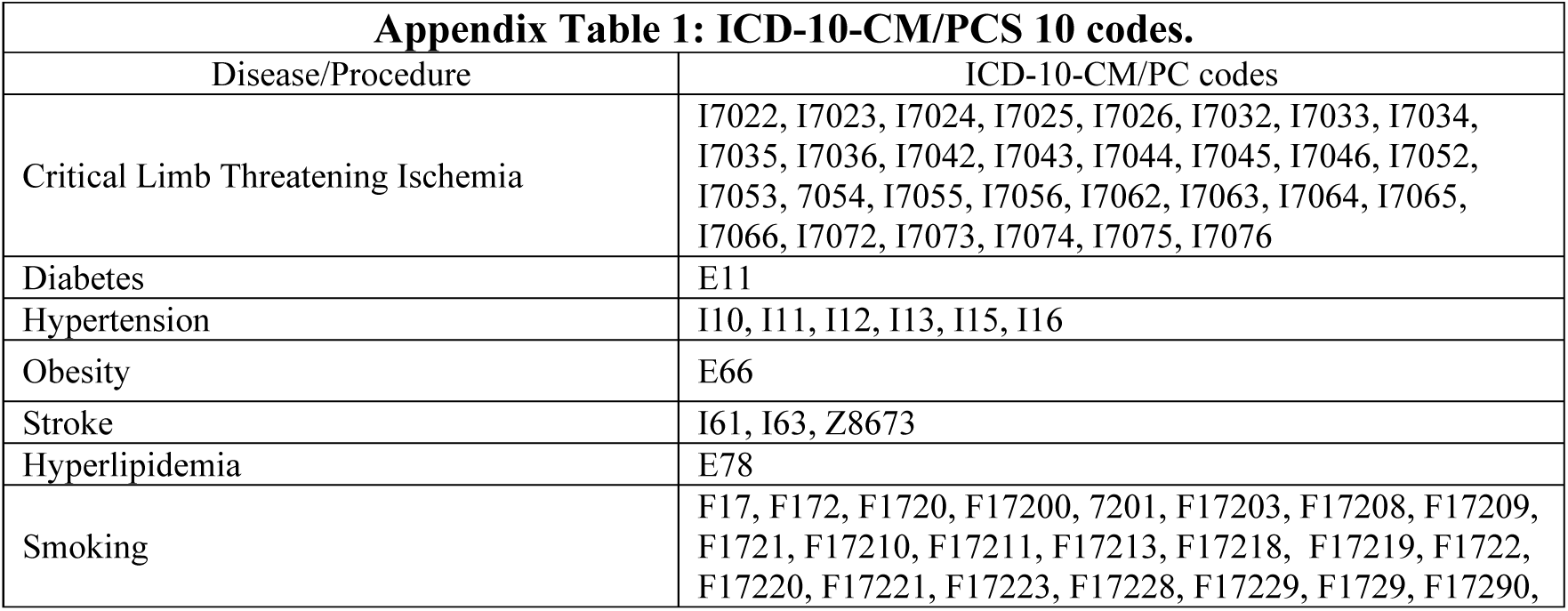
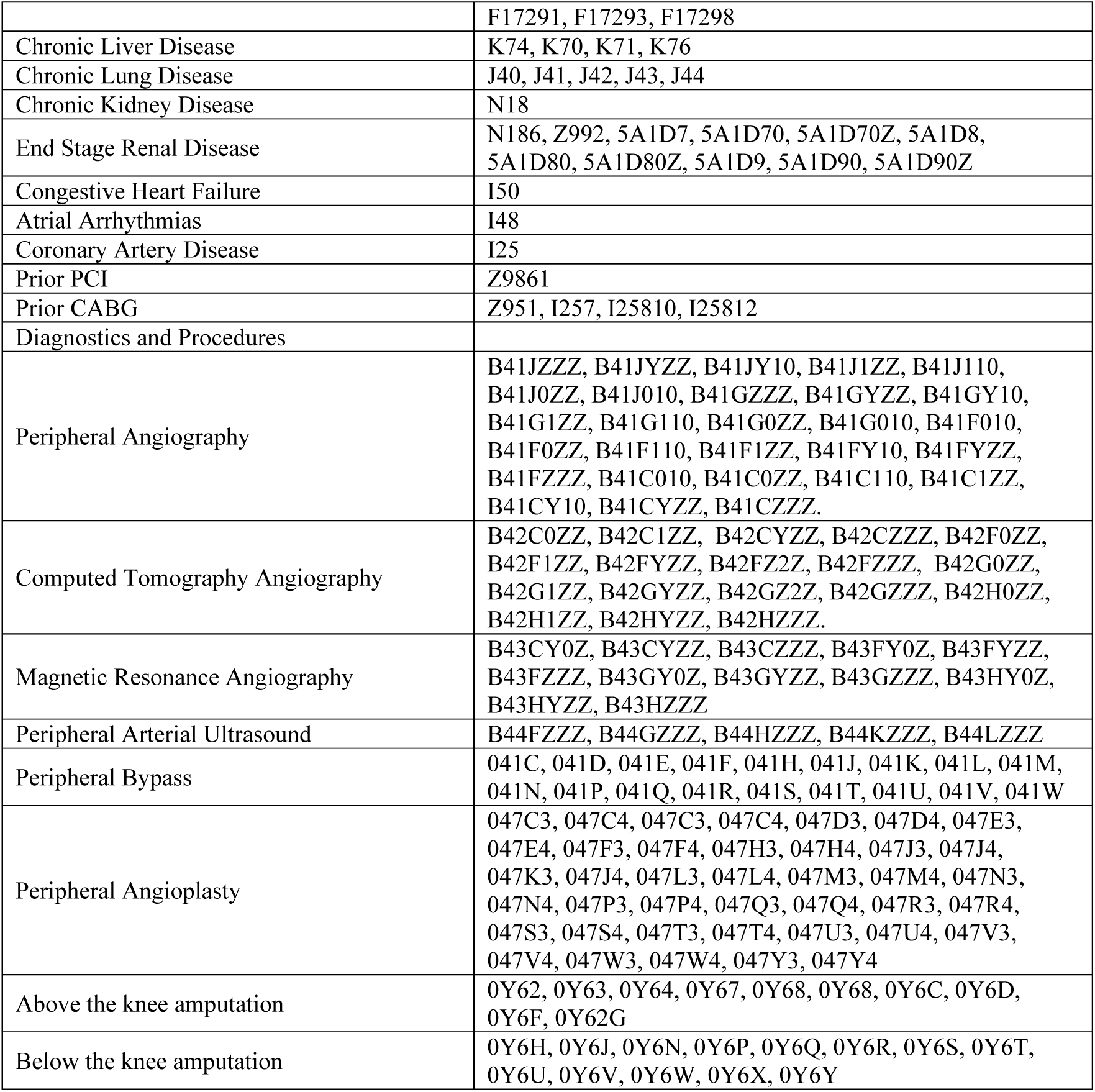

